# Overview of prognostic factors in adult glioma; a 10-year experience at a single institution

**DOI:** 10.1101/2024.03.22.24304679

**Authors:** Amir Barzegar Behrooz, Hadi Darzi Ramandi, Hamid Latifi-Navid, Payam Peymani, Rahil Tarharoudi, Nasrin Momeni, Mohammad Mehdi Sabaghpour Azarian, Sherif Eltonsy, Ahmad Pour-Rashidi, Saeid Ghavami

## Abstract

**Objective:** The most prevalent central nervous system (CNS) neoplasm arising from glial cells is glioma, which diffusely invades brain tissue. Among gliomas, Glioblastoma (GBM) is a glioma of the highest grade and associated with a grim prognosis, with a median overall survival of 15 months with and 3-4 months without therapy. We examined how clinical variables and molecular profiles may have affected overall survival (OS) at Sina Hospital in Tehran over the past ten years.

**Methods:** A retrospective study was conducted at Sina Hospital in Tehran, Iran, and examined patients ≥ 11 years with confirmed glioma diagnoses between 2012 and 2020. We evaluated the correlation between OS in GBM patients and sociodemographic as well as clinical factors, including age, gender, extent of tumor resection, tumor location, chemo/radiotherapy, and molecular profiling based on IDH1, MGMT, TERT, and EGFR status. Kaplan-Meier and multivariate Cox regression models were used to assess patient survival.

**Results:** Following a comprehensive evaluation of medical records, 186 patients were enrolled in the study. The median OS was 20 months, with a 2-year survival rate of 62.5%. Among the 132 patients with available IDH measurements, 105 (79.5%) exhibited IDH1 wild-type tumors. Of the 132 patients with assessed MGMT methylation, 94 (71.2%) had MGMT methylated tumors. TERT promoter methylation was detected in 112 out of 132 cases (84.8%), while no methylation was observed in 20 cases (15.2%). Analyses using multivariable models revealed that age at histological grade (*P* < 0.0001), adjuvant radiotherapy (*P* < 0.014), IDH1 status (*P* < 0.026), and TERT promoter status (*P* < 0.030) were independently associated with OS.

**Conclusion:** The findings of this study demonstrate that patients with higher tumor histological grades who had received adjuvant radiotherapy, exhibited IDH1 mutations, or presented with TERT promoter mutations, experienced improved OS. The median survival time was 20 months, with 15.6% and 46.9% of patients surviving at 12 and 24 months, respectively.

## Introduction

Gliomas are primary brain lesions involving cerebral structures without well-defined boundaries and constitute the most prevalent central nervous system (CNS) neoplasms. Glioblastoma (GBM) is a glioma of the highest grade with, has a very dismal prognosis. Gliomas can develop at any age, but are most common in older adults ^1, 2^. Patients with GBM under 70 years of age showed a median lifespan of around 14.6 months, even with the best current standard of treatment, which includes adjuvant chemotherapy with temozolomide (TMZ) and chemoradiotherapy after tumor excision. Population-based research indicates that chances of survival decline with age ^3, 4^.

Considering most GBM patients pass away from the illness in under a year and almost none survive long-term, GBM tumors have attracted a lot of interest in the research community^5^. Even after extensive surgery, concomitant radiation, adjuvant TMZ, and rigorous treatment, the median survival period for adult patients is still only around ten months, which may get up to 14 months with combination treatment and radiation; just 3-5% of patients live longer than three to five years after diagnosis ^6–8^. In addition to patient characteristics like age and gender, biological variables such as O^6^-methylguanine-DNA methyltransferase (MGMT) promoter methylation status, 1p/19q deletion, and [ isocitrate dehydrogenase 1 (IDH1) gene mutation status may impact disease susceptibility and progression ^9^, which makes predicting GBM survival a challenging task.

Age at diagnosis, surgical resection, tumor grade, chemo/radiotherapy, and genetic profiling affect glioma survival. This study evaluated the characteristics of glioma patients treated at Sina Hospital in Tehran over the past decade. We highlighted and discussed critical issues such as the importance of molecular profiling in predicting OS, the association of glioma survival with prognostically favorable clinical factors, and future research priorities for this patient population.

## Materials and methods

### Population characteristics and study design

This retrospective study involved patients aged 11 years and older hospitalized at Sina Hospital in Tehran, Iran, from March 2012 to September 2020. The Ethics and Research Committee of Tehran University of Medical Science, Neurosurgical Department of Sina Hospital (IR.TUMS.SINAHOSPITAL.REC.1399.111) approved this project. Included patients had a confirmed diagnosis of glioma based on histological examination. Various data points were collected, including age, gender, extent of tumor resection, tumor location, chemo/radiotherapy details, and genomic profiling information such as IDH1 status, MGMT methylation, telomerase reverse transcriptase (TERT) promoter mutation, and epidermal growth factor receptor (EGFR) amplification. Data were obtained by examining patient hospital records and the use of a data collection form. The classification of all tumors was performed according to the WHO classification system. The male-to-female ratio and average age at diagnosis were calculated for each histological subtype. To maintain ethical standards, patient information was extracted from archived records using pseudonyms to ensure the confidentiality of participants. This study does not include any patient-specific information. Patients who underwent emergency operations and those with a Karnofsky Performance Status (KPS) score below 70 and/or a history of psychiatric diseases were excluded (please see **Figure 1** for further details).

**Figure 1.**
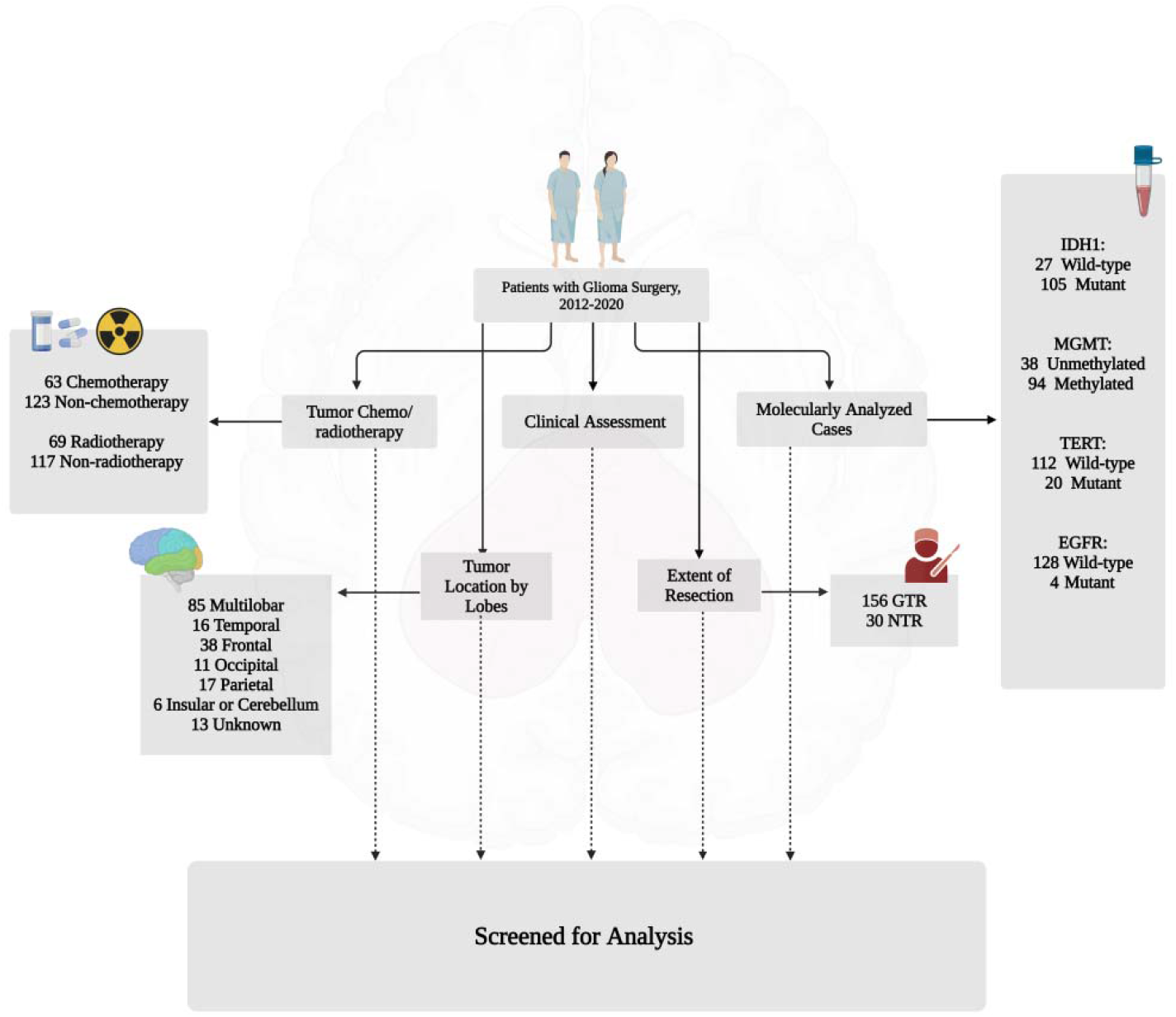
Study design and flow chart of patient selection (Created with BioRender.com).

### Genetic profile

A Qiagen ffpe (formalin-fixed paraffin-embedded) kit was used for sample extraction, Master ABI for PCR, and Qiagen q24 qangen Stanford University for pyrosequencing.

### Statistical analysis

The primary focus of our study was to assess the impact of specific clinical variables and molecular profiles on overall survival (OS), defined as the duration from the day of tumor diagnosis to the date of death from any cause. To gain insights into the characteristics of the patients, we conducted a thorough descriptive analysis. Descriptive statistics were employed to summarize continuous measures, including the number of observed values, median, standard deviation, median, and range. To examine the differences between subgroups, categorized data were compared using the Chi-square (χ^2^) test. Furthermore, we employed the two-tailed Student’s t-test to analyze the age distribution comparison. The reverse Kaplan-Meier method was utilized to estimate the median survival time. Survival curves were generated using Kaplan-Meier estimates, and differences between the curves were analyzed using the log-rank test. A two-tailed *p-*value < 0.05 was considered statistically significant. We used Cox proportional hazards regression analysis to evaluate the impact of measured variables on patient survival in multivariate adjusted models, using a stepwise Wald backward selection procedure. All statistical analyses were performed using *R* software version 4.1.0.

## Results

A complete flow diagram of patient selection is provided in **Figure 1**. This study investigated the association between OS and various factors, including gender, age, distinct tumor grades, the extent of resection, multiple surgeries, radio- and chemotherapy received, mutational profiles in IDH1 and TERT, EGFR alterations, and methylation of the MGMT promoter. Among the cohort of 186 patients meeting the inclusion criteria, a combined treatment approach incorporating both radiation therapy and chemotherapy was administered to 58 patients. Additionally, 11 patients received radiation therapy as a standalone treatment, while 5 patients exclusively underwent chemotherapy.

### Association between glioma grades and parameters studied

Figure 2 provides an overview of the prevalence of genetic and epigenetic alterations identified in the 186 glioma patients with available follow-up. A detailed description of the clinical characteristics and molecular profiling observed in the patients can be found in **Table 1**. Patients were grouped based on the degree of glioma. The median age of patients with grade II, III, and IV tumors was 40.0, 44.5, and 45.5 years, respectively. This patient cohort with newly diagnosed glioma was comprised of 121 (65.1%) males and 65 (34.9%) females, as shown in Figure 3. The χ^2^ value was estimated to compare the expected ratios with the observed ratios separately for sex groups, age groups, tumor grade, radiotherapy, and chemotherapy across different tumor grades. Both men and women demonstrated an increase in mortality with an increasing degree of glioma (grade IV) (Figure 4A). An analysis of patient age and mortality rates revealed that in both age groups (age < 50 and age ≥ 50) the mortality rate significantly higher in patients with grade IV glioma compared to the other groups (Figure 4B). Mortality rate of patients with grade IV tumors in both the unilobar and multilobar groups was significantly higher than in patients with grade II and III glioma (Figure 4C). Only patients with grade IV tumors that underwent chemotherapy showed significantly lower mortality rates and higher survival than patients who did not (Figure 4D). Similarly, patients with grade IV tumors subjected to radiotherapy showed a higher survival rate compared to those not treated (Figure 4E); both chemo- and radiotherapy appeared to improve survival in grade III patients as well, although these effects did not reach statistical significance (Figures 4D, E).

**Figure 2.**
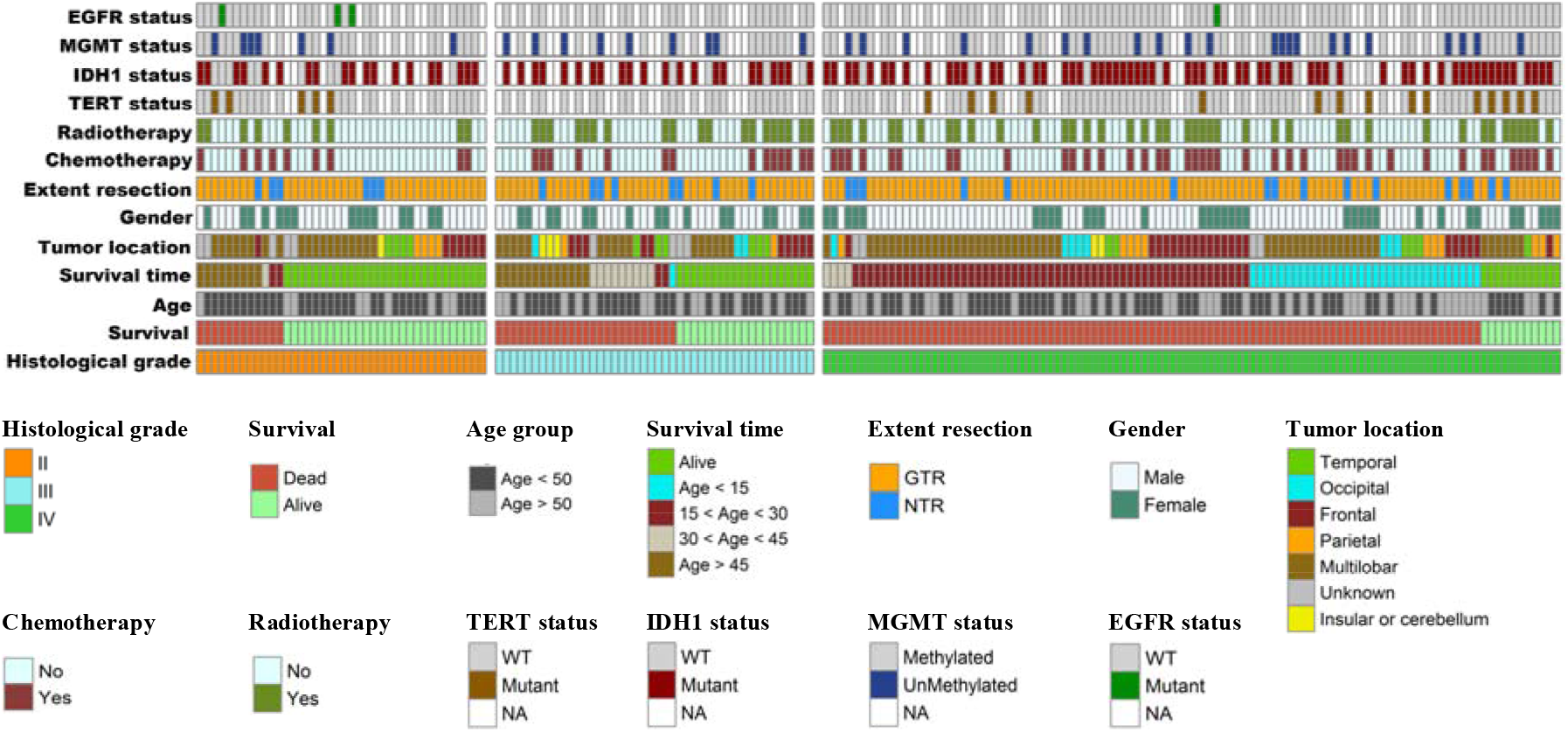
The distribution of molecular and clinical characteristics among the included glioma patient population. Molecular classification was arranged by the three histological types of gliomas (grades II, III, and IV)

**Figure 3.**
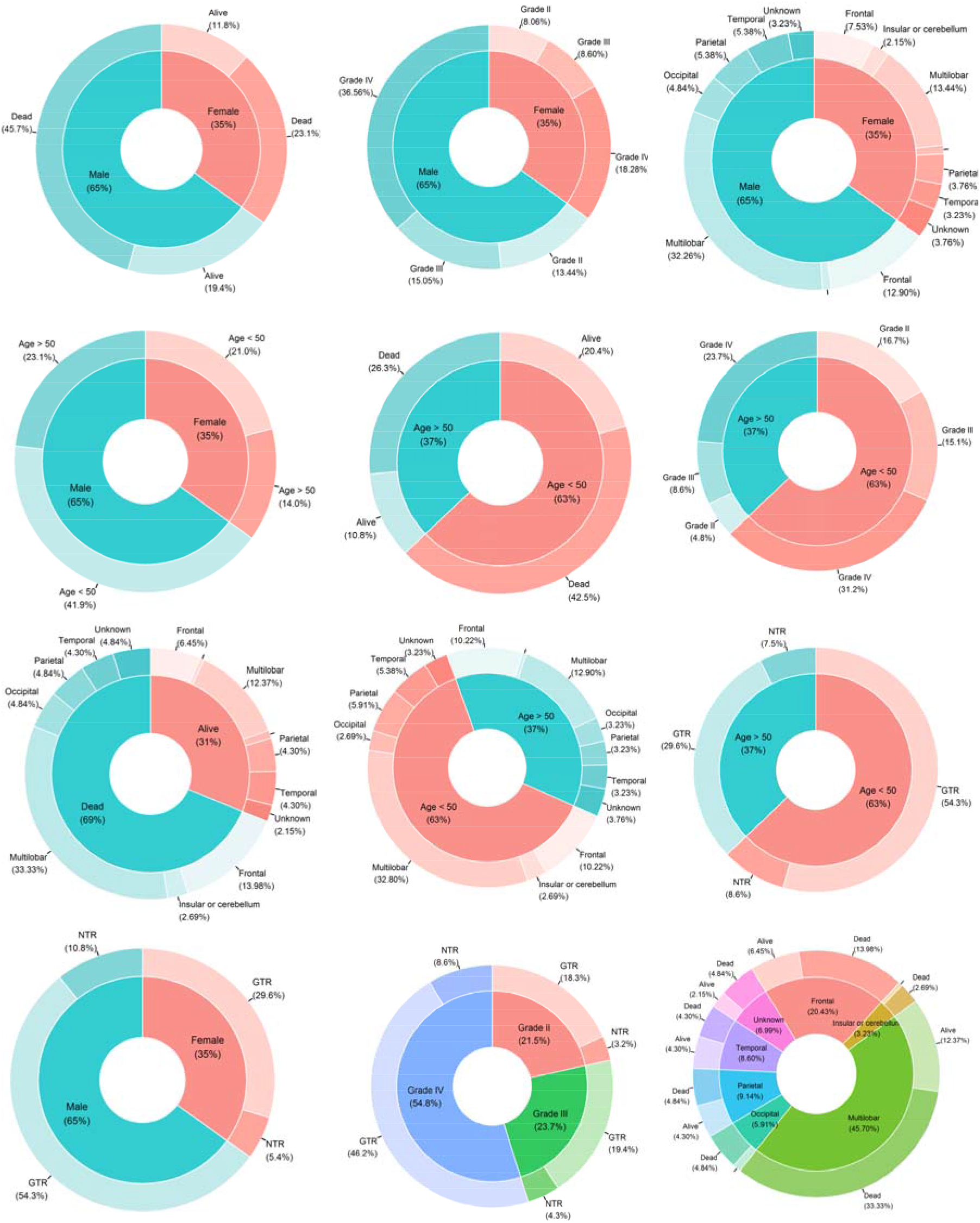
Pie charts showing the frequency of specific characteristics among the 186 patients of the studied glioma cohort. The area of each segment is proportional to the number of patients

**Figure 4.**
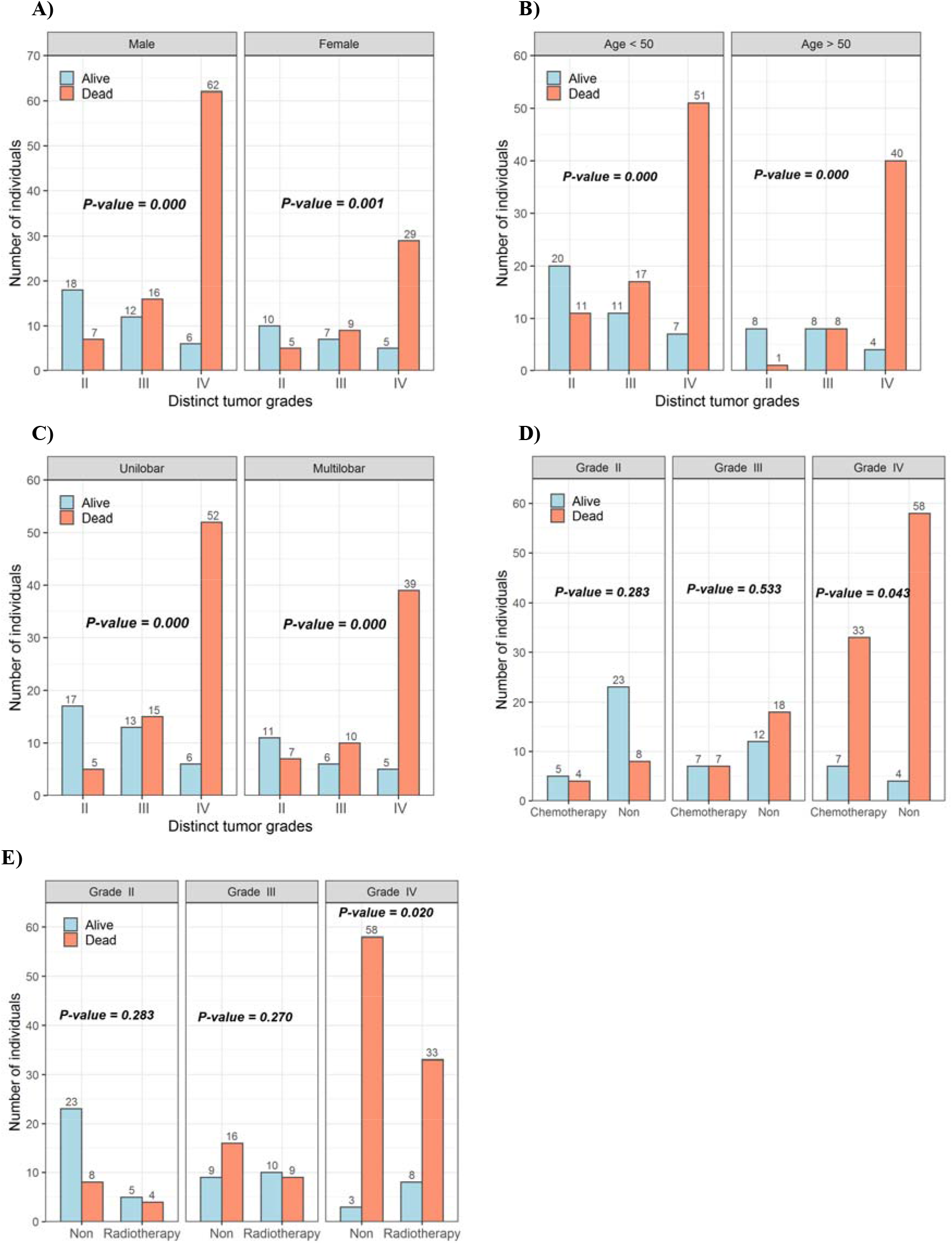
The ratios of different grades of glioma in the examined groups are shown in (**A**) gender *vs.* survival, (**B**) age groups *vs.* survival, (**C**) glioma *vs.* survival, (**D**) chemotherapy *vs.* survival, and (**E**) adjuvant radiotherapy *vs.* survival. The Chi-square test was used to compare the groups

**Table 1.** Characteristics of the 186 patients with glioma in this study.

| Characteristics | Cases | % |
| --- | --- | --- |
| Age group |  |  |
| < 20 years | 8 | 4.3 |
| 20-50 years | 109 | 58.6 |
| > 50 years | 69 | 37.1 |
| Median age (range); years | 44.5 (11 – 81) |  |
| Gender |  |  |
| Male | 121 | 65.1 |
| Female | 65 | 34.9 |
| Distinct tumor grades |  |  |
| Grade II | 40 | 21.5 |
| Grade III | 44 | 23.7 |
| Grade IV | 102 | 54.8 |
| Extent of resection (EOR) |  |  |
| Gross-total resection (GTR) | 156 | 83.9 |
| Near-total resection (NTR) | 30 | 16.1 |
| Tumor location by lobe |  |  |
| Multilobar | 85 | 45.7 |
| Temporal | 16 | 8.6 |
| Frontal | 38 | 20.4 |
| Occipital | 11 | 5.9 |
| Parietal | 17 | 9.1 |
| Insular or cerebellum | 6 | 3.2 |
| Unknown | 13 | 7.0 |
| Survival |  |  |
| Dead | 128 | 68.8 |
| Alive | 58 | 31.2 |
| Survival ≤ 12 | 20 | 15.6 |
| 12 < Survival ≤ 24 | 60 | 46.9 |
| 24 < Survival ≤ 36 | 18 | 14.1 |
| Survival ≥ 36 | 30 | 23.4 |
| Multiple surgeries |  |  |
| Yes | 62 | 33.3 |
| No | 124 | 66.7 |
| Anesthesia |  |  |
| Awake | 22 | 12.6 |
| General | 152 | 87.4 |
| Chemotherapy |  |  |
| Yes | 63 | 33.9 |
| No | 123 | 66.1 |
| Adjuvant radiotherapy |  |  |
| Yes | 69 | 37.1 |
| No | 117 | 62.9 |
| Radio/chemo therapy |  |  |
| Yes | 58 | 31.2 |
| No | 128 | 68.8 |
| Type 2 diabetes mellitus |  |  |
| Yes | 14 | 7.5 |
| No | 172 | 92.5 |

**Table 2.** The distribution of IDH1, MGMT, TERT, and EGFR mutations among different histological grades of glioma in 132 patients with available genetic profiling.

| Molecular assay | Different grades of glioma tumor |  |  | Total |
| --- | --- | --- | --- | --- |
|  | Grade II (28) | Grade III (26) | Grade IV (78) |  |
| IDH1 status |  |  |  |  |
| Wild type | 9 | 4 | 14 | 27 |
| Mutant | 19 | 22 | 64 | 105 |
| MGMT methylation status |  |  |  |  |
| Methylated | 21 | 17 | 56 | 94 |
| Non-methylated | 7 | 9 | 22 | 38 |
| TERT promoter status |  |  |  |  |
| Wild type | 23 | 26 | 63 | 112 |
| Mutant | 5 | 0 | 15 | 20 |
| EGFR amplification status |  |  |  |  |
| Wild type | 25 | 26 | 77 | 128 |
| Mutant | 3 | 0 | 1 | 4 |

### IDH1 and EGFR mutation allele frequencies

In this study, the IDH1 and EGFR status was evaluated in tissue samples from each patient (Figure 2), and the frequencies of IDH1 and EGFR mutations were determined (**Tables 2**). It should be noted that genetic profile information was available for 132 out of the total 186 patients included in the study. Among the grade II gliomas (n=28), 19 cases (67.8%) exhibited IDH1 mutations, while 3 patients (10.7%) had EGFR mutations. In grade III gliomas (n=26), 22 cases (84.6%) showed IDH1 mutations, and no EGFR mutations were observed. In grade IV gliomas (n=78), 64 patients (82.0%) had IDH1 mutations, and 1 patient (1.2%) had an EGFR mutation. Among the 26 grade III glioma samples, 4 cases (15.3%) had wild-type IDH1, and 26 cases (100%) had wild-type EGFR. On the other hand, in the 78 grade IV glioma cases, 14 cases (17.9%) had wild-type IDH1, and 77 cases (98.7%) had wild-type EGFR. It is worth noting that the frequency of IDH1 mutations was higher compared to EGFR mutations in grade II, III, and IV gliomas. Specifically, we observed only 3, 0, and 1 mutation(s) in the EGFR gene among grade II, III, and IV gliomas, respectively.

### Allele frequencies of TERT and MGMT genes and clinical response in patients

In the study cohort of 132 cases, the IDH1, MGMT, TERT, and EGFR status were evaluated successfully. Among these cases, 105 (79.5%) exhibited IDH1 mutations, 20 (15.1%) had TERT mutations, 94 (71.2%) were classified as MGMT-methylated tumors, and four (3%) showed EGFR mutations. MGMT methylation rates were lower (6 patients) in patients with IDH1 wild-type tumors and higher (88 patients) in patients with IDH1 mutant tumors. Of the 186 patients with treatment recorded, 63 (33.9%) and 69 (37.1%) received TMZ and radiotherapy, respectively. As of the time of final data collection, 58 patients (31.2%) were still alive or lost to follow-up.

In this study of 173 patients, tumors in the frontal lobe were the most prevalent (95 patients), followed by tumors in the temporal lobe (76 patients), parietal lobe (68 patients), occipital lobe (29 patients), and insular cortex or cerebellum (19 patients). The most common tumor combinations were observed in the frontal + temporal (18 patients), parietal + frontal (15 patients) and parietal + frontal + temporal (15 patients) lobes (Figure 5A). None of the patients had tumors in all four brain lobes simultaneously. Occipital lobe tumors tended to manifest as isolated cases in glioma patients, with 11 out of 29 cases showing no signs of tumors in other brain lobes (**Figure 3 and 5A**). Anatomically, the location of tumors was primarily multilobar (85 of 173 patients), with a subset of patients demonstrating frontal (21.9%, n = 38) and temporal (9.2%, n = 16) gliomas. Other tumor locations included the parietal lobe (9.8%, n = 17), insular cortex or cerebellum (3.5%, n = 6), and occipital lobe (6.4%, n = 11) (Figure 5A). The tumor location was unknown in 13 patients due to unavailable clinical follow-up and imaging data.

**Figure 5.**
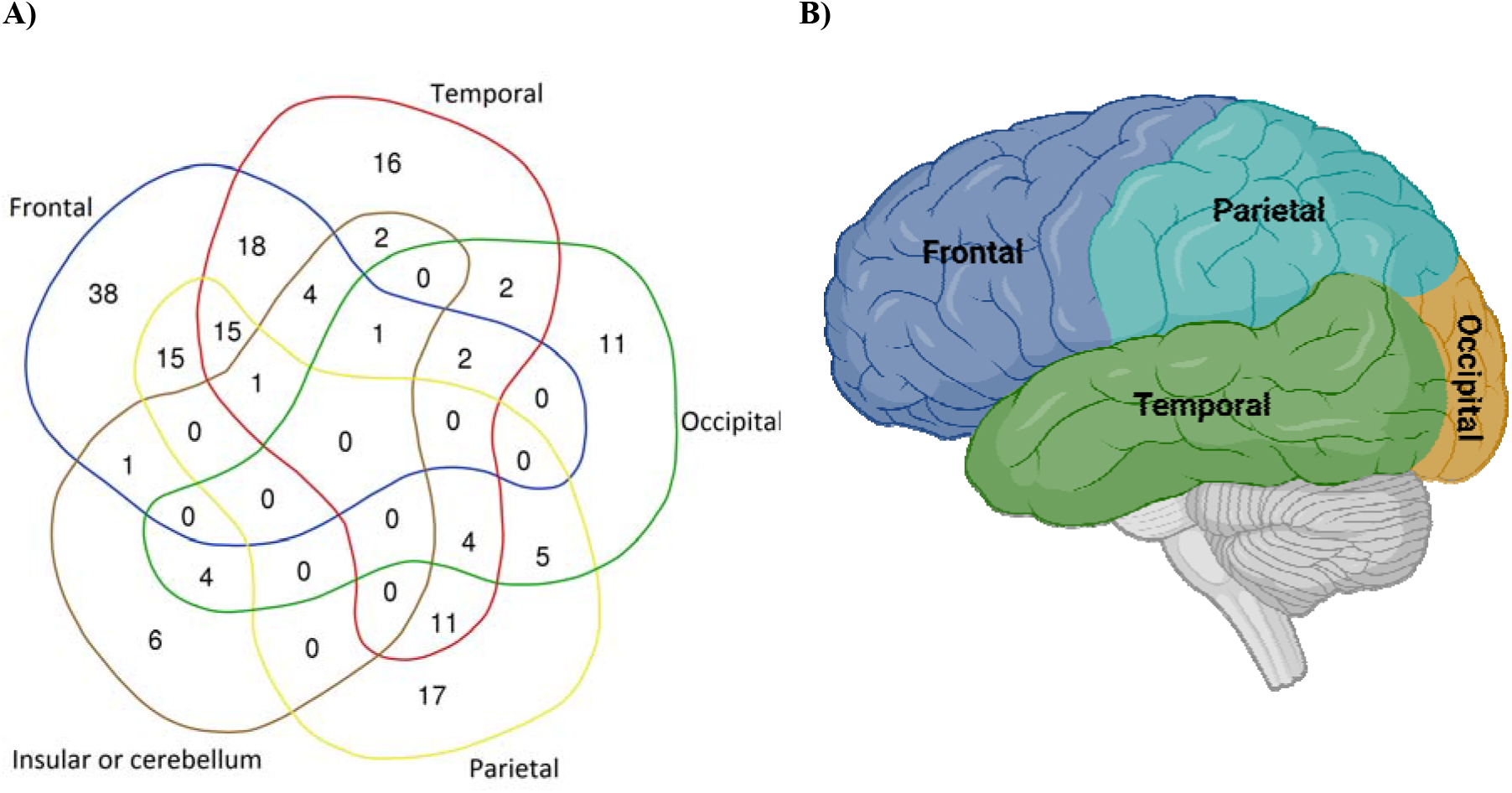
**(A)** Venn diagram illustrating the distribution of 173 patients with tumors in the “Temporal”,” Frontal”,” Occipital”,” Parietal” and “Insular or cerebellum” lobes of the brain. **(B)** The lobes of brain (Created with BioRender.com).

The alluvial plot (Figure 6) shows the distribution and relationships between survival rate, tumor grade, TERT genetic profile, and adjuvant radiotherapy. A considerable number of patients with tumor grade IV (n = 102) had a high mortality rate (90%). Furthermore, the chart illustrates that patients who received adjuvant radiotherapy had a significantly lower mortality rate. The association between the TERT genetic profile and survival rate further underscores the relevance of genetic characteristics in prognostic evaluations.

**Figure 6.**
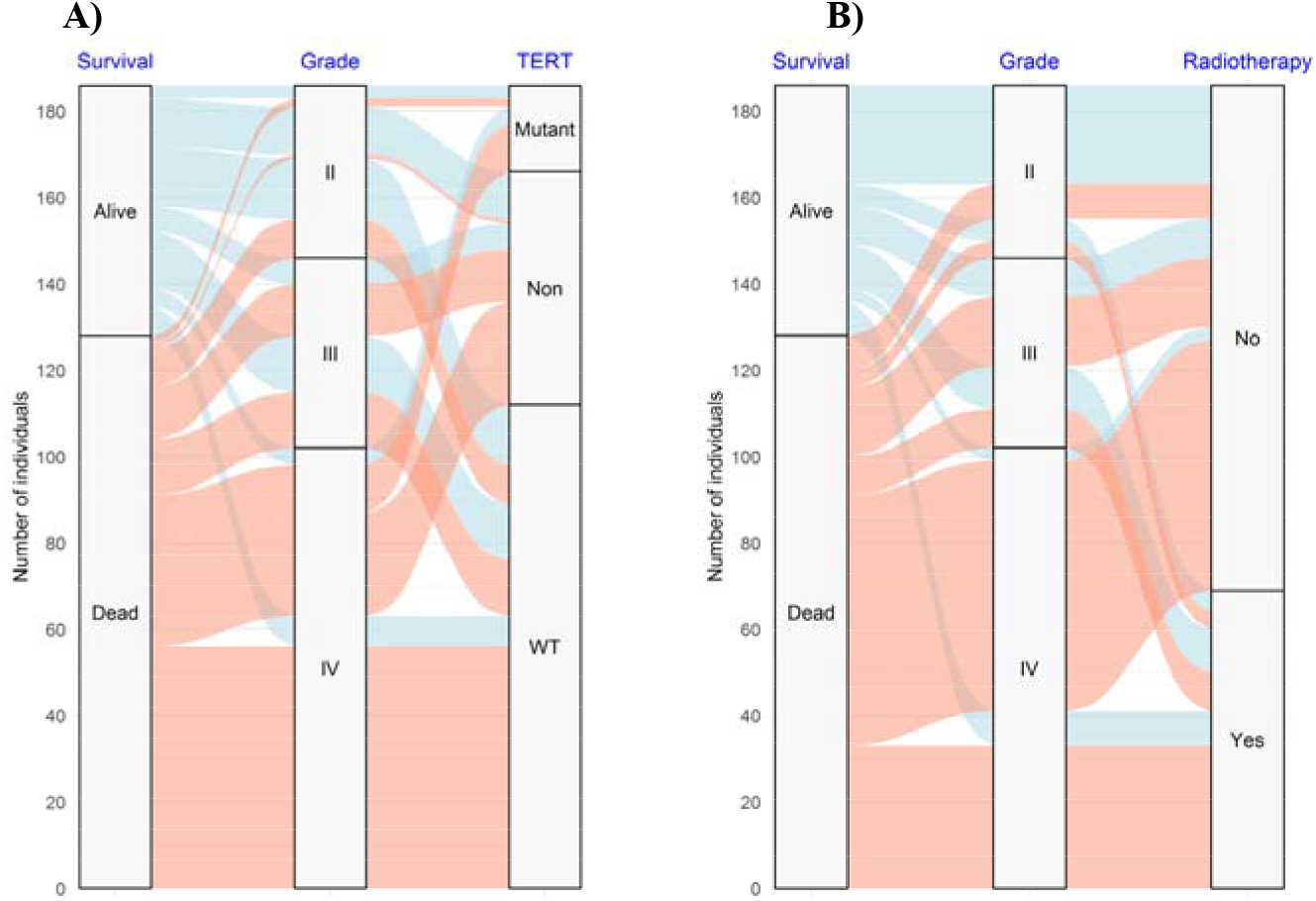
The alluvial plot showing the distribution of patients, survival subgroup, and histological grade in relation to (**A**) TERT alleles and (**B**) adjuvant radiotherapy

### Survival analysis

Results of multivariate Cox regression analyses for clinical characteristics and molecular profiles, including IDH1, TERT, and MGMT, are presented in **Table 3**. In multivariate models, the histological grade of the tumor (HR, 5.70; 95% CI, 3.54-9.18; *P* < 0.0001), adjuvant radiotherapy (HR, 0.41; 95% CI, 0.19-0.83; *P* < 0.014), IDH status (HR for IDH1 mutant status, 2.05; 95%CI, 1.07-4.32; *P* < 0.026), TERT status (HR for unmethylated status, 2.15; 95% CI, 1.08-3.88; *P* < 0.030) were significantly associated with OS (**Table 3**).

**Table 3.** Prognostic univariate and multivariate Cox regression analyses in glioma patients.

| Characteristic | Patients<br>number | Univariate<br>*p*-value | Multivariate |  |  |  |
| --- | --- | --- | --- | --- | --- | --- |
|  |  |  | *p*-value | HR | 95% CI for HR |  |
|  |  |  |  |  | Lower | Upper |
| Age, per year (<50 vs >50) | 128 | 0.022* | 0.0534 | 1.0128 | 0.9998 | 1.026 |
| Gender (Male vs Female) | 128 | 0.26 | 0.4336 | 1.1675 | 0.7923 | 1.7204 |
| Histological grade (grades II, III, and IV) | 128 | $2.9 \times 10^{-13}$* | $7.67 \times 10^{-13}$* | **5.7041** | **3.5431** | **9.1829** |
| Extent of resection (GTR vs NTR) | 128 | 0.55 | 0.6865 | 1.1092 | 0.6705 | 1.835 |
| Glioma (unilobar vs multilobar) | 128 | 0.84 | 0.7637 | 0.9381 | 0.6185 | 1.4228 |
| Chemotherapy (Yes vs No) | 128 | 0.76 | 0.2542 | 1.4972 | 0.7481 | 2.9963 |
| Adjuvant radiotherapy (Yes vs No) | 128 | 0.0047* | 0.0142* | **0.4017** | **0.1938** | **0.8326** |
| IDH1 status (WT vs Mutant) | 91 | 0.83 | 0.0262* | **2.0562** | **1.0749** | **4.317** |
| TERT mutation (WT vs Mutant) | 91 | $2.6 \times 10^{-5}$* | 0.0305* | **2.1542** | **1.0891** | **3.882** |
| MGMT methylation (WT vs Mutant) | 91 | 0.49 | 0.4238 | 0.7986 | 0.4604 | 1.385 |

\* $P < 0.05$ .

### Factors associated with overall survival

The median survival of patients was 20 months (600 days; Figure 7A), with 15.6% and 46.9% of patients alive at 12 and 24 months, respectively. Nineteen patients (14.8%) survived for longer than 48 months (4 years) and were considered long-term survivors. Interestingly, the four patients who were still alive when the dataset was finalized (OS > 50, 53, 65, and 71 months) exhibited mutated IDH1 and methylated MGMT. Initially, we assessed whether any genetic alterations were associated with survival through univariate and multivariate analyses (**Table 3**). A more stringent multivariate analysis, which only incorporates parameters with *P* > 0.05 in the univariate analyses, revealed the same parameters were independent prognostic factors. Glioma patients with IDH1 mutations had a median OS of 21 months compared to 17.5 months for those without an IDH1 mutation (Figure 7G). Methylation of MGMT was associated with a median OS of 20.5 months compared to 19 months in non-methylated cases (Figure 7I). Patients with a TERT promoter mutation exhibited a median OS of 16 months, whereas patients with a non-mutated TERT promoter had a median OS of 21 months (Figure 7H). Notably, patients with solely an IDH1 mutation exhibited the most favorable survival, with a median survival of 21 months. Subsequently, patients with both an IDH1 mutation and MGMT methylation experienced a slightly lower median survival of 20 months. In contrast, patients lacking either an IDH1 mutation or MGMT methylation had the shortest median survival of 16 months. It is worth noting that the molecular profile analysis, specifically regarding wild-type IDH1 and MGMT methylation, encompassed a limited sample size of only three patients who exhibited survival durations of 18, 50, and 62 months, respectively. Furthermore, we conducted an assessment of the prognostic significance of histological grade and clinical characteristics. Among the histological subgroups, Grades II and III demonstrated a more favorable prognosis, with median OS durations of 51 and 46 months, respectively. Conversely, Grade IV was associated with an extremely poor outcome, with an OS of 18 months (Figure 7B). Additionally, patients under the age of 50 displayed a more favorable prognosis, with a median OS of 23 months, compared to the subgroup aged 50 years and above, which exhibited a median OS of 19 months (Figure 7D).

**Figure 7.**
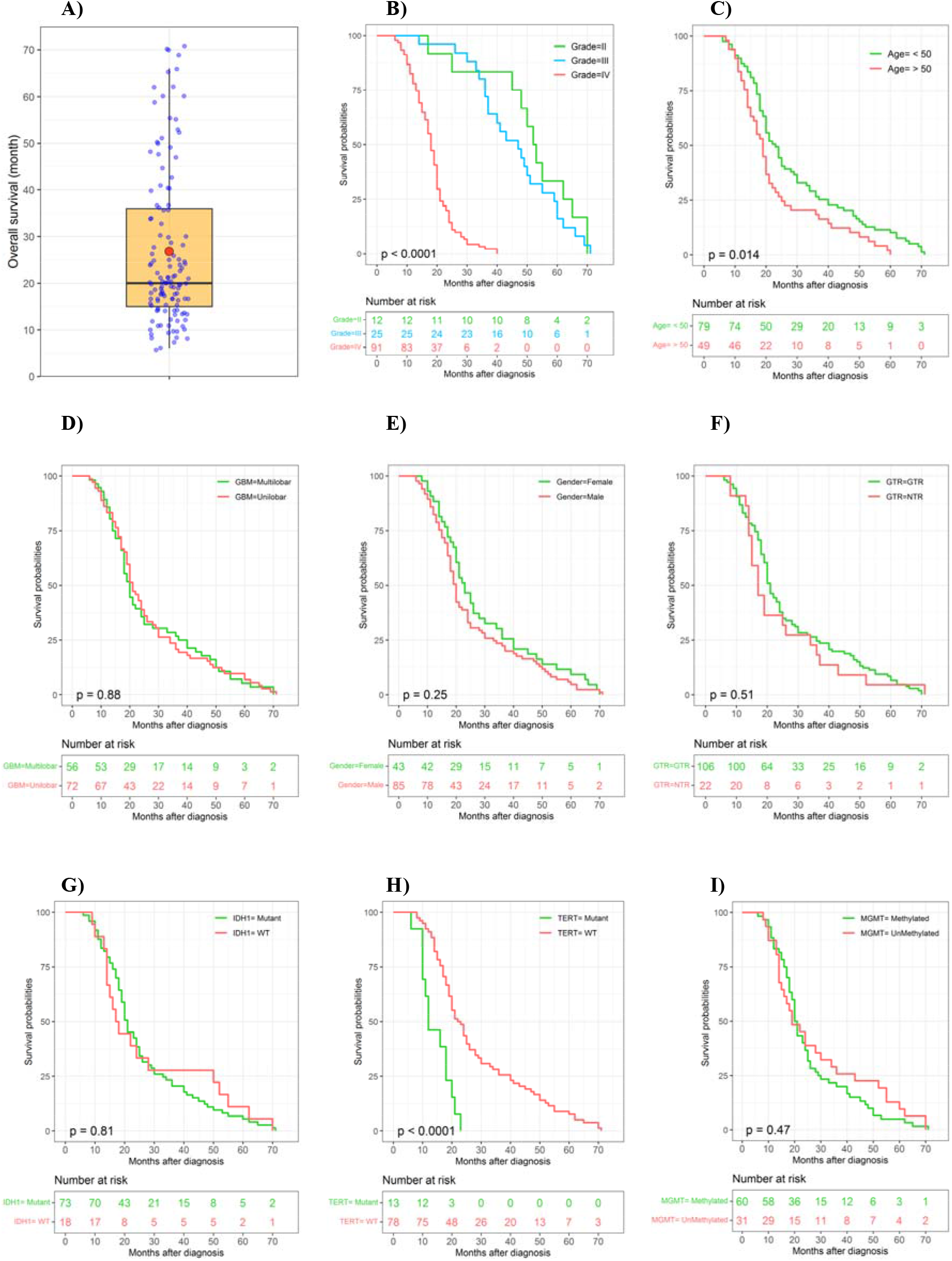
The distribution of glioma patient survival and Kaplan-Meier curves for overall survival are shown for 8 risk groups. **(A)** The box plot illustrates the overall survival (months) of glioma patients. Kaplan-Meier curves show survival probabilities in relation to **(B)** the histological grade, **(C)** age groups, **(D)** uni- and multilobar glioma, **(E)** gender, **(F)** extent of resection, **(G)** IDH1 status, (H) TERT status, and (I) MGMT status.

## Discussion

Our results showed a difference of 40, 44.5, and 45.5 years in the median age of patients with grade II, III, and IV tumors, respectively. Gliomas were more common in males than in females, and the death rate increased with glioma grade, with the highest mortality in grade IV patients. GBM is the most prevalent malignant brain tumor in adults, accounting for 54% of all cases. The incidence of GBM increases with age, and the development of illnesses, especially those affecting the CNS, is more likely among older people. Wild-type IDH GBM is the most common aggressive primary brain tumor in adults, with an average diagnostic age of 68–70 years, and progresses rapidly. Elderly (65 and older) GBM patients may have a worse post-treatment survival rate due to age-related changes, such as diminished immune system function and persistent neuroinflammation ^10, 11^. A negative correlation exists between the age at which GBM is identified and prognosis. The 10-year relative survival rate declines from 15.2% to 5.7% between 20-44 and 45-54 years of age, respectively ^12, 13^.

The results of the current study show that glioma patients with IDH1 mutations (IDH1mt) had a median OS of 21 months versus 17.5 months for those without an IDH1 mutation. In addition, methylation of the MGMT (MGMTmet) was associated with a median OS of 20.5 months versus 19 months in patients with a non-methylated. The combination of IDH1 mutations and MGMT methylation status is a more accurate predictor of survival in glioblastoma compared to either IDH1 or MGMT alone. Glioblastoma patients were categorized into 3 distinct genotypes based on the genetic and epigenetic characteristics of IDH1 and MGMT: GBM patients with IDH1mt/MGMTmet had the longest survival, followed by patients with IDH1mt/MGMTunmet or IDH1wt/MGMTmet, and patients with IDH1wt/MGMTunmet had the shortest survival^14^. Improved prognosis has been linked to tumor molecular characteristics such as MGMT promoter methylation and IDH1 mutation ^15–17^. Thus, MGMT methylation positively impacts survival and responsiveness to TMZ therapy^15^. In addition, IDH and MGMT co-methylation is linked to a better prognosis and predicts the response to chemotherapy and surgical resection ^16^.

Surgical resection, radiation, and TMZ chemotherapy constitute the standard treatment for GBM. Despite the intensive nature of this therapy, the tumor reoccurs within 7 to 10 months following surgery in 75-90% of GBM patients ^18–20^. A study utilizing molecular testing revealed distinct genetic profiles among GBM survivors. Notably, mutations in the IDH genes (IDH1 and IDH2) and methylation of the MGMT promoter were identified as two significant factors associated with a more favorable response to standard clinical care ^21^. Patients with mutant IDH1/2 GBM exhibited superior outcomes to those with wild-type IDH tumors, with a survival of 14 and 42 months, respectively ^22^. Literature suggests a strong correlation between TERT gene polymorphisms and an increased risk of glioma in patients ^23^. TERT mutations are also linked to biomarkers such as IDH1, 1p19q, TP53, and EGFR ^24^.

Gliomas are more prevalent in men than in women. The tumors of glioblastoma patients showed huge genetic sex differences linked to survival. According to a study on differentially expressed genes in the tumor clusters, survival in men was impacted the most by genes that govern cell division. Regarding survival in females, integrin gene expression was the most critical mechanism for tumor dissemination [23]. Other work indicated that male patients had the lowest cancer-specific survival (CSS) rates throughout localized cancer stages and various age groupings, which was confirmed by stratified analysis ^25, 26^.

One study reported that women with IDH1-mutant tumors were concentrated in the group with the most favorable prognosis, while in males; the mutations were spread out across all groups. Considering IDH1 mutations have been linked to higher survival in glioblastoma patients, the longest-surviving female cluster is consistent with this theory. However, this was not the case for men ^25^. Our present study indicates that patients with a mutation in TERT had a median OS of 16 months compared to 21 months in those without a TERT mutation. In addition, MGMT methylation status was shown to affect the prognostic value of a TERT promoter mutation. Only TERT-mutated GBM with MGMT methylation may respond to TMZ; therefore, TMZ may not benefit all patients with MGMT-methylated GBM ^27^. Long-term follow-up of patients with TERT promoter mutant GBMs showed a high correlation between the prognosis and the presence of multifocal/distant lesions. There was a associated between EGFR amp/gain, CDKN2A deletion, and PTEN loss, as well as a negative correlation between CDK4 and TP53 deletion for the TERTp mutation ^28^. EGFR mutations have been demonstrated to be effective prognostic indicators of OS in IDH-wildtype GBM patients. Moreover, there is data to suggest that EGFR amplification is evident in high-grade gliomas (25%). EGFR amplification was also observed to be limited to IDH wild-type (26%) and TERT mutant (27%) gliomas, occurring irrespective of MGMT promoter methylation status and being mutually exclusive with 1p/19q co-deletion (LOH) ^29^. The functional connection between EGFR and p53 in GBM is intriguing. Thus, EGFR has been shown to reduce the activity of wild-type p53 by increasing the interaction between DNA-PKcs and p53. Either EGFR or DNA-PKc knockdown enhanced the transcriptional activity of wild-type p53 due to a reduced interaction between p53 and DNA-PKcs. These results revealed a unique non-canonical regulatory axis between EGFR and wild-type p53 in GBM, with unexpected biological functions ^30^.

We observed an apparent difference (although not statistically significant) in the OS of gross (GTR) versus near total resection (NTR) glioma patients, which is consistent with findings by Abdelfath et al. ^31^. GTR appears to be more beneficial than subtotal resection (STR) in extending the life of elderly individuals with high-grade glioma ^32^. Aggressive surgical resection should be considered for older GBM patients, especially those with relatively low KPS. Intraoperative magnetic resonance imaging (ioMRI) does not seem to provide any significant advantage over intraoperative ultrasonography (ioUS) in experienced hands in this population. Still, it may significantly prolong the duration of surgery, which is a modifiable prognostic factor that affects care ^33^. Other work showed a correlation between maximal tumor excision and OS in all categories of glioblastoma patients. Additionally, maximal resection of non–contrast-enhanced (NCE) tumors was associated with longer OS in younger patients, independent of IDH status, and in patients with IDH–wild-type glioblastoma, regardless of the methylation status of the promoter region of the DNA repair enzyme O^6^-methylguanine-DNA methyltransferase ^34^.

Within our cohort, the anatomical tumor location was generally confined to a multi-lobe, primarily multilobar disease (45.6%), with some patients demonstrating tumor growth in the frontal (20.4%) and temporal (8.6%) lobes. Other tumor locations included the parietal lobe (9.1%), insular cortex or cerebellum (3.2%), and occipital lobe (5.9%). Most glioblastomas develop in the periventricular white matter areas close to the subventricular zone. MGMT promoter methylation tumors are more prevalent in the left temporal lobe, particularly in patients with GBM, an IDH1 mutant tumor, tumors with the proneural gene expression subtype, or frontal lobe tumors missing PTEN deletion. IDH1 mutation-associated MGMT-methylated tumors tended to manifest in the left frontal lobe, whereas EGFR-amplified and EGFR variant 3-expressing tumors occurred most frequently in the left temporal lobe. A comparable area in the left temporal lobe was associated with excellent radiochemotherapy response and improved survival^35^. In another study, tumors in the right occipitotemporal periventricular white matter were substantially related to poor survival in both training and test cohorts and had a greater tumor volume than tumors in other regions. Right parietal tumors were associated with hypoxia pathway enrichment and platelet-derived growth factor receptor (PDGFRA) amplification, deeming these processes appropriate subgroup-specific treatment targets. Additionally, central tumor placement was associated with a worse prognosis. In elderly individuals, the distance from the center of the third ventricle to the contrast-enhancing tumor border may be a practical prognostic indicator ^35–37^.

Our study recognizes certain limitations, including the utilization of data from a single center and an underpowered analysis for detecting differences among patient subgroups. The scarcity of data on comorbidities such as lung infections, renal disorders, seizures, high blood pressure, and paresthesia emphasizes the necessity for more extensive and standardized data collection. Moreover, we did not adjust for multiple comparisons. However, the study’s strength lies in its meticulous examination of factors associated with glioma prognosis and treatment response, presenting valuable insights for clinical decision-making. Additionally, we highlighted the synergistic impact of specific genetic profiles and radiotherapy, identifying potential targets for personalized treatment strategies. Despite the acknowledged limitations, this study offers a comprehensive analysis of crucial factors influencing glioma prognosis and treatment response.

## Conclusion

The present study aimed to investigate the impact of clinical and molecular factors on the overall survival of glioblastoma patients. Multivariate Cox regression analysis revealed that histological tumor grade, adjuvant radiotherapy, IDH status, and TERT promoter status were significantly associated with overall survival. Notably, patients with IDH1 mutations and TERT promoter mutations exhibited longer survival. Factors such as extensive tumor removal, smaller tumor size, and prompt initiation of radiation therapy after surgery were associated with favorable prognoses. Patients with higher tumor grades had poorer outcomes, while those who received adjuvant radiotherapy showed improved survival. In conclusion, this study highlights the importance of molecular and clinical characteristics in predicting overall survival in glioblastoma patients and provides valuable insights for personalized treatment strategies.

## Ethic

The Ethics and Research Committee of Tehran University of Medical Science, Neurosurgical Department of Sina Hospital (IR.TUMS.SINAHOSPITAL.REC.1399.111) approved this study.

## Acknowledgement

We appreciate Sina Hospital for providing the required testing facilities and equipment for the study.

## Author contribution

**Amir Barzegar Behrooz:** drafting & editing of the manuscript with input from all authors. **Hadi Darzi Ramandi:** contributed to data analysis and interpretation, and drafting & editing of the manuscript. **Hamid Latifi-Navid:** reviewing & editing of the manuscript. **Payam Peymani:** reviewing & editing of the manuscript. **Rahil Tarharoudi, Nasrin Momeni and Mohammad Mehdi Sabaghpour Azarian:** assisted in data curation. **Sherif Eltonsy:** Reviewing & editing of the manuscript. **Ahmad Pour-Rashidi:** supervised the project and contributed to the writing, reviewing, and editing. **Saeid Ghavami:** supervision of the project, resources, and final draft. All authors approved the final document as submitted.

## Data availability statement

The authors confirm that the data supporting the findings of this study are available within the article.

## References

(1) Dunn, G. P.; Rinne, M. L.; Wykosky, J.; Genovese, G.; Quayle, S. N.; Dunn, I. F.; Agarwalla, P. K.; Chheda, M. G.; Campos, B.; Wang, A.;, et al. Emerging insights into the molecular and cellular basis of glioblastoma. Genes Dev 2012, 26 (8), 756–784. DOI: 10.1101/gad.187922.112 From NLM.

(2) Nahid Dadashzadeh Asl1a, T. S. a., Meisam Haghmoradi 3, Ahmad Pour-Rashidi 4,; Mohammad Mehdi Sabaghpour Azarian 5, H. H., *, Vahid Hosseinpour 7, Amir; Barzegar Behrooz 8. Prevalence of Central Nervous System Tumors in Iran: A Systematic and Meta-analysis. Latin American Journal of Pharmacy 2023, 42 (10), 8–17.

(3) Minniti, G.; Lombardi, G.; Paolini, S. Glioblastoma in Elderly Patients: Current Management and Future Perspectives. Cancers (Basel) 2019, 11 (3). DOI: 10.3390/cancers11030336 From NLM.

(4) Weller, M.; Wick, W.; Aldape, K.; Brada, M.; Berger, M.; Pfister, S. M.; Nishikawa, R.; Rosenthal, M.; Wen, P. Y.; Stupp, R.;, et al. Glioma. Nature Reviews Disease Primers 2015, 1 (1), 15017. DOI: 10.1038/nrdp.2015.17.

(5) Zhang, Y. H.; Li, Z.; Zeng, T.; Pan, X.; Chen, L.; Liu, D.; Li, H.; Huang, T.; Cai, Y. D. Distinguishing Glioblastoma Subtypes by Methylation Signatures. Front Genet 2020, 11, 604336. DOI: 10.3389/fgene.2020.604336 From NLM.

(6) Dubrow, R.; Darefsky, A. S. Demographic variation in incidence of adult glioma by subtype, United States, 1992-2007. *BMC Cancer* 2011, *11*, 325. DOI: 10.1186/1471-2407-11-325 From NLM.

(7) van den Bent, M. J.; Geurts, M.; French, P. J.; Smits, M.; Capper, D.; Bromberg, J. E. C.; Chang, S. M. Primary brain tumours in adults. Lancet 2023, 402 (10412), 1564–1579. DOI: 10.1016/s0140-6736(23)01054-1 From NLM.

(8) Mohammed, S.; Dinesan, M.; Ajayakumar, T. Survival and quality of life analysis in glioblastoma multiforme with adjuvant chemoradiotherapy: a retrospective study. Rep Pract Oncol Radiother 2022, 27 (6), 1026–1036. DOI: 10.5603/RPOR.a2022.0113 From NLM.

(9) Baid, U.; Rane, S. U.; Talbar, S.; Gupta, S.; Thakur, M. H.; Moiyadi, A.; Mahajan, A. Overall Survival Prediction in Glioblastoma With Radiomic Features Using Machine Learning. Front Comput Neurosci 2020, 14, 61. DOI: 10.3389/fncom.2020.00061 From NLM.

(10) Miller, K. D.; Ostrom, Q. T.; Kruchko, C.; Patil, N.; Tihan, T.; Cioffi, G.; Fuchs, H. E.; Waite, K. A.; Jemal, A.; Siegel, R. L.; et al. Brain and other central nervous system tumor statistics, 2021. CA Cancer J Clin 2021, 71 (5), 381–406. DOI: 10.3322/caac.21693 From NLM.

(11) Kim, M.; Ladomersky, E.; Mozny, A.; Kocherginsky, M.; O’Shea, K.; Reinstein, Z. Z.; Zhai, L.; Bell, A.; Lauing, K. L.; Bollu, L.;, et al. Glioblastoma as an age-related neurological disorder in adults. Neurooncol Adv 2021, 3 (1), vdab125. DOI: 10.1093/noajnl/vdab125 From NLM.

(12) Rabab’h, O.; Al-Ramadan, A.; Shah, J.; Lopez-Negrete, H.; Gharaibeh, A. Twenty Years After Glioblastoma Multiforme Diagnosis: A Case of Long-Term Survival. Cureus 2021, 13 (6), e16061. DOI: 10.7759/cureus.16061 From NLM.

(13) Ostrom, Q. T.; Gittleman, H.; Truitt, G.; Boscia, A.; Kruchko, C.; Barnholtz-Sloan, J. S. CBTRUS Statistical Report: Primary Brain and Other Central Nervous System Tumors Diagnosed in the United States in 2011-2015. Neuro Oncol 2018, 20 (suppl_4), iv1-iv86. DOI: 10.1093/neuonc/noy131 From NLM.

(14) Molenaar, R. J.; Verbaan, D.; Lamba, S.; Zanon, C.; Jeuken, J. W. M.; Boots-Sprenger, S. H. E.; Wesseling, P.; Hulsebos, T. J. M.; Troost, D.; van Tilborg, A. A.;, et al. The combination of IDH1 mutations and MGMT methylation status predicts survival in glioblastoma better than either IDH1 or MGMT alone. Neuro-Oncology 2014, 16 (9), 1263–1273. DOI: 10.1093/neuonc/nou005 (acccessed 3/4/2024).

(15) Louis, D. N.; Perry, A.; Reifenberger, G.; von Deimling, A.; Figarella-Branger, D.; Cavenee, W. K.; Ohgaki, H.; Wiestler, O. D.; Kleihues, P.; Ellison, D. W. The 2016 World Health Organization Classification of Tumors of the Central Nervous System: a summary. Acta Neuropathol 2016, 131 (6), 803–820. DOI: 10.1007/s00401-016-1545-1 From NLM.

(16) Molenaar, R. J.; Verbaan, D.; Lamba, S.; Zanon, C.; Jeuken, J. W.; Boots-Sprenger, S. H.; Wesseling, P.; Hulsebos, T. J.; Troost, D.; van Tilborg, A. A.;, et al. The combination of IDH1 mutations and MGMT methylation status predicts survival in glioblastoma better than either IDH1 or MGMT alone. Neuro Oncol 2014, 16 (9), 1263–1273. DOI: 10.1093/neuonc/nou005 From NLM.

(17) Hegi, M. E.; Diserens, A. C.; Gorlia, T.; Hamou, M. F.; de Tribolet, N.; Weller, M.; Kros, J. M.; Hainfellner, J. A.; Mason, W.; Mariani, L.;, et al. MGMT gene silencing and benefit from temozolomide in glioblastoma. N Engl J Med 2005, 352 (10), 997–1003. DOI: 10.1056/NEJMoa043331 From NLM.

(18) Ho, V. K.; Reijneveld, J. C.; Enting, R. H.; Bienfait, H. P.; Robe, P.; Baumert, B. G.; Visser, O. Changing incidence and improved survival of gliomas. Eur J Cancer 2014, 50 (13), 2309–2318. DOI: 10.1016/j.ejca.2014.05.019 From NLM.

(19) Aldape, K.; Zadeh, G.; Mansouri, S.; Reifenberger, G.; von Deimling, A. Glioblastoma: pathology, molecular mechanisms and markers. Acta Neuropathol 2015, 129 (6), 829–848. DOI: 10.1007/s00401-015-1432-1 From NLM.

(20) Omuro, A.; DeAngelis, L. M. Glioblastoma and other malignant gliomas: a clinical review. Jama 2013, 310 (17), 1842–1850. DOI: 10.1001/jama.2013.280319 From NLM.

(21) Jovčevska, I. Genetic secrets of long-term glioblastoma survivors. Bosn J Basic Med Sci 2019, 19 (2), 116–124. DOI: 10.17305/bjbms.2018.3717 From NLM.

(22) Molinaro, A. M.; Taylor, J. W.; Wiencke, J. K.; Wrensch, M. R. Genetic and molecular epidemiology of adult diffuse glioma. Nat Rev Neurol 2019, 15 (7), 405–417. DOI: 10.1038/s41582-019-0220-2 From NLM.

(23) Yuan, Y.; Qi, C.; Maling, G.; Xiang, W.; Yanhui, L.; Ruofei, L.; Yunhe, M.; Jiewen, L.; Qing, M. TERT mutation in glioma: Frequency, prognosis and risk. J Clin Neurosci 2016, 26, 57–62. DOI: 10.1016/j.jocn.2015.05.066 From NLM.

(24) Powter, B.; Jeffreys, S. A.; Sareen, H.; Cooper, A.; Brungs, D.; Po, J.; Roberts, T.; Koh, E. S.; Scott, K. F.; Sajinovic, M.;, et al. Human TERT promoter mutations as a prognostic biomarker in glioma. J Cancer Res Clin Oncol 2021, 147 (4), 1007–1017. DOI: 10.1007/s00432-021-03536-3 From NLM.

(25) Yang, W.; Warrington, N. M.; Taylor, S. J.; Whitmire, P.; Carrasco, E.; Singleton, K. W.; Wu, N.; Lathia, J. D.; Berens, M. E.; Kim, A. H.;, et al. Sex differences in GBM revealed by analysis of patient imaging, transcriptome, and survival data. Sci Transl Med 2019, 11 (473). DOI: 10.1126/scitranslmed.aao5253 From NLM.

(26) Tian, M.; Ma, W.; Chen, Y.; Yu, Y.; Zhu, D.; Shi, J.; Zhang, Y. Impact of gender on the survival of patients with glioblastoma. Biosci Rep 2018, 38 (6). DOI: 10.1042/bsr20180752 From NLM.

(27) Vuong, H. G.; Nguyen, T. Q.; Ngo, T. N. M.; Nguyen, H. C.; Fung, K. M.; Dunn, I. F. The interaction between TERT promoter mutation and MGMT promoter methylation on overall survival of glioma patients: a meta-analysis. BMC Cancer 2020, 20 (1), 897. DOI: 10.1186/s12885-020-07364-5 From NLM.

(28) Kikuchi, Z.; Shibahara, I.; Yamaki, T.; Yoshioka, E.; Shofuda, T.; Ohe, R.; Matsuda, K. I.; Saito, R.; Kanamori, M.; Kanemura, Y.;, et al. TERT promoter mutation associated with multifocal phenotype and poor prognosis in patients with IDH wild-type glioblastoma. Neurooncol Adv 2020, 2 (1), vdaa114. DOI: 10.1093/noajnl/vdaa114 From NLM.

(29) Hölzl, D.; Hutarew, G.; Zellinger, B.; Alinger-Scharinger, B.; Schlicker, H. U.; Schwartz, C.; Sotlar, K.; Kraus, T. F. J. EGFR Amplification Is a Phenomenon of IDH Wildtype and TERT Mutated High-Grade Glioma: An Integrated Analysis Using Fluorescence In Situ Hybridization and DNA Methylome Profiling. Biomedicines 2022, 10 (4). DOI: 10.3390/biomedicines10040794 From NLM.

(30) Ding, J.; Li, X.; Khan, S.; Zhang, C.; Gao, F.; Sen, S.; Wasylishen, A. R.; Zhao, Y.; Lozano, G.; Koul, D.;, et al. EGFR suppresses p53 function by promoting p53 binding to DNA-PKcs: a noncanonical regulatory axis between EGFR and wild-type p53 in glioblastoma. Neuro Oncol 2022, 24 (10), 1712–1725. DOI: 10.1093/neuonc/noac105 From NLM.

(31) AbdelFatah, M. A. R.; Kotb, A.; Said, M. A.; Abouelmaaty, E. M. H. Impact of extent of resection of newly diagnosed glioblastomas on survival: a meta-analysis. Egyptian Journal of Neurosurgery 2022, 37 (1), 3. DOI: 10.1186/s41984-022-00145-1.

(32) Han, Q.; Liang, H.; Cheng, P.; Yang, H.; Zhao, P. Gross Total vs. Subtotal Resection on Survival Outcomes in Elderly Patients With High-Grade Glioma: A Systematic Review and Meta-Analysis. Front Oncol 2020, 10, 151. DOI: 10.3389/fonc.2020.00151 From NLM.

(33) Barak, T.; Vetsa, S.; Nadar, A.; Jin, L.; Gupte, T. P.; Fomchenko, E. I.; Miyagishima, D. F.; Yalcin, K.; Vasandani, S.; Gorelick, E.;, et al. Surgical strategies for older patients with glioblastoma. J Neurooncol 2021, 155 (3), 255–264. DOI: 10.1007/s11060-021-03862-z From NLM.

(34) Molinaro, A. M.; Hervey-Jumper, S.; Morshed, R. A.; Young, J.; Han, S. J.; Chunduru, P.; Zhang, Y.; Phillips, J. J.; Shai, A.; Lafontaine, M.;, et al. Association of Maximal Extent of Resection of Contrast-Enhanced and Non-Contrast-Enhanced Tumor With Survival Within Molecular Subgroups of Patients With Newly Diagnosed Glioblastoma. JAMA Oncol 2020, 6 (4), 495–503. DOI: 10.1001/jamaoncol.2019.6143 From NLM.

(35) Ellingson, B. M.; Lai, A.; Harris, R. J.; Selfridge, J. M.; Yong, W. H.; Das, K.; Pope, W. B.; Nghiemphu, P. L.; Vinters, H. V.; Liau, L. M.;, et al. Probabilistic radiographic atlas of glioblastoma phenotypes. AJNR Am J Neuroradiol 2013, 34 (3), 533–540. DOI: 10.3174/ajnr.A3253 From NLM.

(36) Fyllingen, E. H.; Bø, L. E.; Reinertsen, I.; Jakola, A. S.; Sagberg, L. M.; Berntsen, E. M.; Salvesen, Ø.; Solheim, O. Survival of glioblastoma in relation to tumor location: a statistical tumor atlas of a population-based cohort. Acta Neurochir (Wien) 2021, 163 (7), 1895–1905. DOI: 10.1007/s00701-021-04802-6 From NLM.

(37) Liu, T. T.; Achrol, A. S.; Mitchell, L. A.; Du, W. A.; Loya, J. J.; Rodriguez, S. A.; Feroze, A.; Westbroek, E. M.; Yeom, K. W.; Stuart, J. M.;, et al. Computational Identification of Tumor Anatomic Location Associated with Survival in 2 Large Cohorts of Human Primary Glioblastomas. AJNR Am J Neuroradiol 2016, 37 (4), 621–628. DOI: 10.3174/ajnr.A4631 From NLM.

